# Quantitative CT Scoring for Local COPD Severity

**DOI:** 10.1101/2025.04.09.25324951

**Authors:** Wassim W. Labaki, Sundaresh Ram, Ali Namvar, Alexander J. Bell, Benjamin A. Hoff, Ella A. Kazerooni, Stefanie Galban, Fernando J. Martinez, Charles R. Hatt, Susan Murray, Evgeny M. Mirkes, Alexander N. Gorban, Andrei Zinovyev, MeiLan K. Han, Craig J. Galban

**Author notes:** Corresponding Author: Craig J. Galban, PhD, Department of Radiology, University of Michigan 109 Zina Pitcher Place, BSRB A506, Ann Arbor, MI 48109-2200, USA.

## Abstract

Chronic obstructive pulmonary disease (COPD) is complex, and its course is difficult to predict due to its diverse pathophysiology. Small airway disease (SAD), a key component of COPD and potential target for emerging therapeutics, may be reversible in mild COPD, but left unchecked, may worsen, leading to airway loss and emphysema. The dual nature of SAD complicates clinical management of COPD patients, necessitating more accurate monitoring methods. To meet this need, we developed elastic Parametric Response Mapping (ePRM), a tiered scoring system that classifies local lung volumes by the degree of PRM-derived SAD, normal, and emphysematous tissue. In individuals with or at risk for COPD, we demonstrate that chest CT ePRM can categorize local lung tissue into distinct tiers of disease severity that distinguish between tissue characterized by early reversible SAD and progressive destruction. This level of characterization is crucial to developing personalized treatment strategies for COPD.

## Introduction

Chronic obstructive pulmonary disease (COPD) is the third leading cause of death in the world and a significant source of morbidity for the estimated half billion individuals living with the condition [1–3]. Beyond the diagnostic criterion of irreversible airflow obstruction (currently defined as a post-bronchodilator forced expiratory volume in one second to forced vital capacity ratio (FEV_1_/FVC) < 0.7), COPD is characterized by a complex clinical heterogeneity that is not captured by spirometric measurements alone [4]. Chest computed tomography (CT) provides additional anatomic and functional assessments to help identify COPD phenotypes and understand disease progression [5, 6]. CT assessments can be based on computational techniques such as Parametric Response Mapping (PRM), developed by our group. PRM detects spatially-resolved density changes between co-registered inspiratory and expiratory chest CT acquisitions to distinguish alveolar destruction due to emphysema (PRM^Emph^) from non-emphysematous air trapping, a surrogate measure of small airway disease (SAD) referred to as functional small airway disease (PRM^fSAD^) [7]. Importantly, PRM has been validated as a biomarker of airway and alveolar abnormalities as measured using micro-CT tissue analysis of explanted human lung specimens and is commercially available [8].

PRM analysis of chest CT scans has been performed in several large COPD observational studies, including Genetic Epidemiology of COPD (COPDGene) [9] and the SubPopulations and Intermediate Outcome Measures in COPD Study (SPIROMICS) [10]. Over the past decade, PRM analysis has helped define COPD phenotypes and predict disease progression [11]. It has clearly illustrated the heterogeneity of COPD, since patients with similar severities of airflow obstruction on spirometry can have strikingly different PRM readouts [7]. Additionally, PRM analysis has built on the results of seminal physiologic and histologic studies [12, 13] to show that SAD is an early lesion in COPD pathogenesis and a bridge to irreversible emphysema. For example, COPDGene participants with mild COPD (FEV_1_ ≥ 80% predicted) had on average more than 20% of their total lung volume quantified by PRM^fSAD^ [14]. Further, PRM^fSAD^ has been independently associated with subsequent lung function decline and emphysema progression in individuals with or at risk of COPD [14–16].

Despite these advances, the dual nature of SAD complicates interpretation of PRM-defined fSAD. Current PRM outputs are presented as percentages of total lung volume for each classification (i.e., normal lung, emphysema, fSAD, and parenchymal disease). Although easy to calculate, these measures overlook the spatial variability of SAD, where reversible and irreversible forms may be present. Thus, lung disease depicted by PRM remains incompletely characterized at the local level, but understanding how differences in local PRM composition may relate to specific disease states—specifically, how they may infer progression—is particularly relevant for SAD. We have histologically demonstrated that in severe disease, PRM^fSAD^ is characterized by airway fibrosis and even airway loss [8]. However, we have also seen PRM^fSAD^ revert to normal classification, particularly in patients with milder disease, suggesting that early in disease, the imaging biomarker is picking up reversible processes such as occluding airway mucus plugs or reversible infiltration of immune cells in airway walls [17, 18]. Hence, a better understanding of local lung composition is critical to more precisely phenotype disease and improve our ability to identify patients with local reversible disease for whom more aggressive early intervention or therapeutics may be warranted [19].

We have addressed this limitation through our PRM-based severity scoring system. Referred to as ‘elastic PRM’ or ‘ePRM’, our method uses an elastic principal graphing model to score regional lung volumes based on their PRM composition into five discrete, adjoining tiers (**Fig. 1**). A key component of our approach to analyzing PRM is the ability to not only score local lung tissue but also infer potential pathways for progression from normal lung parenchyma (**Fig. 1**; Tier 0) to fSAD-dominant (**Fig. 1**; Tier 2) or emphysema-dominant (**Fig. 1**; Tier 3) obstructive disease. Although the unbiased elastic principal graphing algorithm is not new, and has been used by our group and others in various clinical and biologic contexts [20–25], its application here is unique. In contrast to standard methods for analyzing PRM^fSAD^, which provide a percent volume of PRM^fSAD^, applying ePRM to 3D PRM data generates a severity scoring map that identifies and quantifies the early onset of disease. Here we demonstrate our ePRM technique in subjects “at-risk” or with confirmed COPD and show how local severity scores advance over time and correlate with lung function over a 5-year period.

**Fig. 1:**
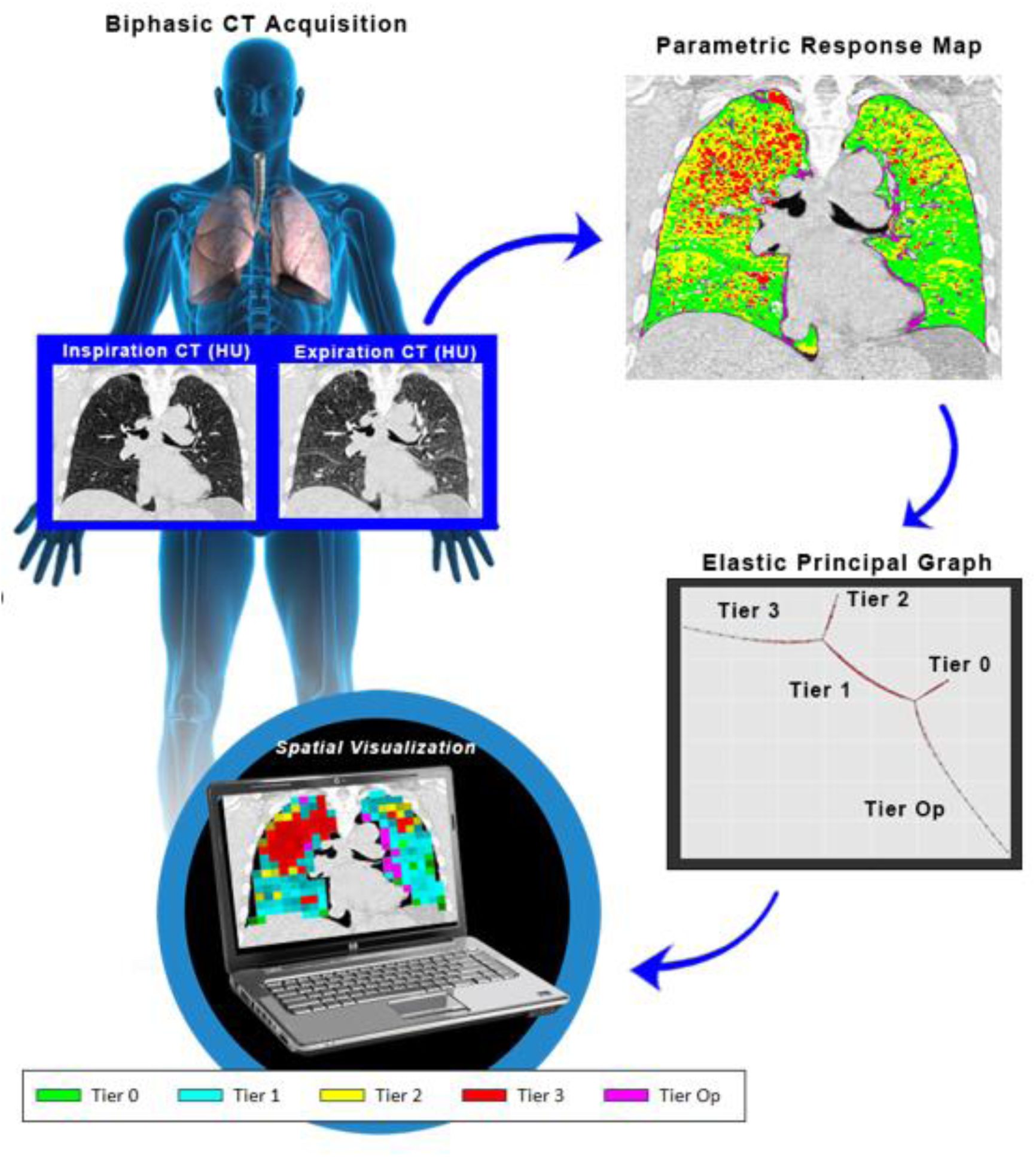
Schematic Diagram of the ePRM workflow. Elastic PRM (ePRM) classifies local PRM composition based on an elastic principal graph that consists of five adjoining clusters that infer progression. Each cluster is classified as a tier severity score based on distinct PRM characteristics. Tiers 0, 2, 3, and Op (terminal segments) are characterized by: elevated PRM^Norm^, elevated PRM^fSAD^ and negligible PRM^Emph^, transition of PRM^fSAD^ to PRM^Emph^, and elevated PRM^PD^ (opacities that result in high attenuation areas), respectively. These tier scores are connected by a single cluster, i.e. bridge, referred to as Tier 1, which is characterized by PRM^Norm^ transitioning to PRM^fSAD^. PRM profiles along trajectories on elastic principal graph are provided in **Supplemental** Fig. 2.

## Results

### Elastic PRM readouts better reflect COPD severity

We first wanted to determine the relationship between ePRM readouts and COPD severity as defined by Global Initiative for Chronic Obstructive Lung Disease (GOLD). Demographic, clinical and high-resolution CT data at baseline and 5-year follow-up were acquired from subjects accrued as part of Phase 1 and Phase 2 of the COPDGene study. Out of the 4,585 subjects evaluated at Phase 2, 3,631 had adequate CT acquisitions for deformable image registration and complete data for longitudinal analyses. Baseline subject demographics, spirometry, exercise capacity and CT metrics, including PRM and ePRM readouts, are provided in **Table 1**. The percent volume of Tier 0 lung decreased with increasing COPD severity, with inverse behavior observed for Tiers 2 and 3. These trends align with percent volumes of PRM^Norm^, PRM^fSAD^ and PRM^Emph^, respectively. The percent volume of Tier 1 showed negligible change with increasing COPD severity. Subjects diagnosed with GOLD 1 COPD had 16±9% (mean ± standard deviation) PRM^fSAD^ and negligible PRM^Emph^. We found for GOLD 1 cases that Tier 1 made up nearly a quarter of the lung volume (23±14%), whereas combined Tiers 2 and 3 contributed on average only 10% of lung volume. In cases with GOLD 4 COPD, the percent volume of PRM^fSAD^ (35±9%) was higher than PRM^Emph^ (26±14%). These results are consistent with previously published works [7, 11]. In contrast, the percent volume of Tier 2 (25±18%) was lower than Tier 3 (43±25%). The mean positions of sub-volumes in Tiers 0, 2 and 3 behaved similarly to the relative volumes of the tiers with increasing COPD severity. Unlike the Tier 1 percent volume, the Tier 1 mean position increased with COPD severity from 0.3±0.08 to 0.51±0.08 in GOLD 1 and 4, respectively. Our ePRM technique suggests that local COPD evolves from Tier 0 to 3, with Tier 1, having negligible COPD dependence, acting as a critical bridge from normal to obstructive lung tissue.

**Table 1:** Subject characteristics at baseline.

| Variable | Non-COPD |  | COPD |  |  |  |
| --- | --- | --- | --- | --- | --- | --- |
|  | PRISm | "At-Risk" | GOLD 1 | GOLD 2 | GOLD 3 | GOLD 4 |
| Participants (N) | 399 (11%) | 1759 (48%) | 320 (9%) | 729 (20%) | 360 (10%) | 64 (2%) |
| Age (yrs) | 58.14 (8.47) | 58.06 (8.36) | 62.24 (8.44) | 62.54 (8.35) | 63.83 (7.86) | 63.41 (6.94) |
| Sex (M/F) | 180/219 | 872/887 | 173/147 | 393/336 | 210/150 | 33/31 |
| BMI (kg/m<sup>2</sup>) | 32.42 (7.38) | 29.2 (5.67) | 27 (4.68) | 28.62 (5.56) | 28.54 (6.13) | 26.56 (5.17) |
| 6MWD (ft) | 1352.24 (356.49) | 1552.04 (353.52) | 1572 (335.77) | 1396.62 (346.62) | 1204.96 (344.16) | 1120.02 (279.94) |
| Smoking Status (f/c) | 196/203 | 925/834 | 164/156 | 402/327 | 236/124 | 50/14 |
| Scanner Make (G/S/P) | 143/14/242 | 630/57/1072 | 104/9/207 | 273/24/432 | 115/15/230 | 25/0/39 |
| Smoking (pack-yrs) | 40.32 (21.18) | 36.97 (20.19) | 44.62 (22.4) | 49.2 (23.85) | 54.68 (25.97) | 54.83 (26.41) |
| **Pulmonary Parameters:** |  |  |  |  |  |  |
| FEV<sub>1</sub> (L) | 2.1 (0.48) | 2.89 (0.68) | 2.67 (0.65) | 1.93 (0.51) | 1.2 (0.29) | 0.72 (0.16) |
| FEV<sub>1</sub> (% predicted) | 71.18 (7.61) | 97.59 (11.53) | 90.98 (8.41) | 65.77 (8.39) | 41.49 (5.59) | 24.98 (3.56) |
| FVC (L) | 2.77 (0.66) | 3.69 (0.9) | 4.11 (0.99) | 3.33 (0.89) | 2.74 (0.75) | 2.35 (0.59) |
| FVC (% predicted) | 72.57 (8.4) | 96.39 (11.65) | 107.22 (11.57) | 86.97 (12.62) | 71.91 (12.91) | 62.02 (11.09) |
| FEV<sub>1</sub>/FVC | 0.76 (0.05) | 0.79 (0.05) | 0.65 (0.04) | 0.58 (0.08) | 0.45 (0.09) | 0.31 (0.05) |
| SGRQ | 26.32 (22.4) | 14.4 (16.31) | 16 (16.24) | 29.02 (21.06) | 40.47 (18.88) | 49.55 (18.5) |
| **Quantitative CT** |  |  |  |  |  |  |
| TLC (L) | 4.73 (1.07) | 5.46 (1.27) | 6.19 (1.47) | 5.83 (1.36) | 6.18 (1.43) | 6.75 (1.14) |
| FRC (L) | 2.63 (0.6) | 2.77 (0.67) | 3.28 (0.84) | 3.5 (0.87) | 4.27 (1.1) | 5.07 (1.02) |
| **Parametric Response Map (%)** |  |  |  |  |  |  |
| PRM<sup>Norm</sup> | 58 (12.39) | 64.32 (10.34) | 58.48 (10.77) | 50.42 (12.55) | 34.91 (12.17) | 23.21 (9.26) |
| PRM<sup>fSAD</sup> | 8.14 (6.11) | 9.03 (7.23) | 15.9 (9.05) | 20.53 (10.16) | 30.81 (9.8) | 35.04 (9.21) |
| PRM<sup>Emph</sup> | 0.64 (1.19) | 0.89 (1.53) | 3.26 (4.15) | 5.61 (7.04) | 13.72 (11.45) | 25.66 (13.93) |
| PRM<sup>PD</sup> | 32.2 (14.56) | 24.17 (10.79) | 19.69 (7.63) | 21.3 (8.62) | 18.69 (7.86) | 14.63 (3.1) |
| **Elastic PRM** |  |  |  |  |  |  |
| **Assignment (%)** |  |  |  |  |  |  |
| Tier 0 | 51 (24) | 65 (21) | 53 (22) | 37 (23) | 14 (15) | 4 (5) |
| Tier 1 | 8 (10) | 10 (11) | 23 (14) | 27 (14) | 29 (12) | 21 (11) |
| Tier 2 | 1 (3) | 2 (6) | 7 (9) | 10 (11) | 21 (15) | 25 (18) |
| Tier 3 | 0 (1) | 0 (1) | 3 (7) | 7 (13) | 22 (22) | 43 (25) |
| Tier Op | 39 (27) | 22 (21) | 15 (14) | 19 (17) | 14 (16) | 7 (6) |
| **Relative Position** |  |  |  |  |  |  |
| Tier 0-p | 0.64 (0.15) | 0.75 (0.11) | 0.73 (0.11) | 0.62 (0.15) | 0.47 (0.16) | 0.34 (0.16) |
| Tier 1-p | 0.24 (0.08) | 0.25 (0.09) | 0.3 (0.08) | 0.33 (0.09) | 0.44 (0.1) | 0.51 (0.08) |
| Tier 2-p | 0.35 (0.34) | 0.42 (0.33) | 0.61 (0.24) | 0.64 (0.2) | 0.71 (0.12) | 0.72 (0.11) |
| Tier 3-p | 0.1 (0.19) | 0.13 (0.19) | 0.25 (0.18) | 0.27 (0.2) | 0.37 (0.18) | 0.51 (0.17) |
| Tier Op-p | 0.24 (0.18) | 0.17 (0.13) | 0.15 (0.09) | 0.18 (0.09) | 0.23 (0.08) | 0.27 (0.07) |

**Note:** Subject characteristics at baseline separated by GOLD status. Continuous and categorical variables are displayed as mean (standard deviation) and counts, respectively. GOLD, Global Initiative for Chronic Obstructive Lung Disease; PRISm, preserved ratio impaired spirometry; "at-risk", at-risk smokers with normal spirometry; Participants, number of participants per GOLD (percentage of total population); Age, (years); Sex, (male/female); BMI, body mass index; 6MWD, 6-minute walking distance in feet; Smoking, smoking status in pack-years; Smoking Status (former smoker/current smoker); Scanner Make (GE/Siemens/Philips); FEV<sub>1</sub>, forced expiratory volume in one second in liters and percent predicted (pp); FVC, forced vital capacity in liters and percent predicted; SGRQ, St. George Respiratory Questionnaire; TLC, CT-based total lung capacity in liter; FRC, functional residual capacity in liter; PRM, parametric response map; Norm, Normal; fSAD, functional small airways disease; Emph, emphysema; PD, parenchymal disease. PRM metrics are percent of total lung volume.

### Local tier scores follow a pattern of progression as identified in a subject

Due to the strong dependence of our ePRM readouts to COPD severity as observed in **Table 1**, we set out to determine if local changes in Tier scores progress in a predictable pattern. Presented in **Fig. 2** are the PRM and ePRM findings for a subject diagnosed with GOLD 2 COPD at baseline who demonstrated a decline in FEV_1_ percent predicted (FEV_1_pp) from 51.4% (GOLD 2) to 41.0% (GOLD 3) over 5 years. We first conducted a visual assessment of local changes in PRM and Tier scores and calculated the distribution of PRM^fSAD^ within each Tier score over 5 years (**Fig. 2a**). At baseline, 54% of the lung volume was comprised of normal lung parenchyma (PRM^Norm^; green voxels), and 27% of PRM^fSAD^ (yellow voxels), with a large consolidation of PRM^fSAD^ in the right lower lung. Of the 27% of PRM^fSAD^ at baseline, about half (52% of all PRM^fSAD^ in the lungs) was associated with Tier 1 and about a third (35% of all PRM^fSAD^ in the lungs) with Tier 2. Consistent with the drop in FEV_1_pp at year 5, the percent volume of PRM^fSAD^ increased by 12% to 39%, with PRM^Emph^ showing a 2% increase to 5%. The distribution of PRM^fSAD^ flipped from baseline values, with the majority of PRM^fSAD^ associated with Tier 2 (57% of all PRM^fSAD^ in the lungs), rather than Tier 1 (32% of all PRM^fSAD^ in the lungs). Overall, the percent volume of lung scored Tier 1 remained unchanged at 38% over the 5-year interval (**Fig. 2b**). In contrast, the percent volume of Tier 2 increased by 21%, from 12% to 33%, and the percent volume of Tier 0 decreased by 22%, from 40% to 18%. While the percent volume of local lung scored Tier 1 (38%) did not change over the time interval, this tier acted as a critical bridge of disease progression, as 52% of lung scored Tier 0 advanced to Tier 1 and 59% of lung scored Tier 1 progressed to Tiers 2 (55%) and 3 (4%). To better understand the role of Tier 1 as a bridge from normal to obstructive lung parenchyma, we evaluated the specific position of local lung on Tier 1 (**Fig. 2c**). To illustrate changes in Tier 1 position, which is a continuous value within the Tier, we stratified local volumes within Tier 1 into four discrete bins (0-0.25, 0.25-0.5, 0.5-0.75, and 0.75-1) based on their position on the Tier 1 segment. Among the sub-volumes in Tier 1 that have position values less than 0.25 at baseline (comprising 43% of sub-volumes), nearly 64% remained in Tier 1, while 33% advanced to Tier 2 at year 5 (**Fig. 2c**). Thirty-three percent of Tier 1 sub-volumes with a position value between 0.25 and 0.5 remained in Tier 1, while 62% advanced to Tier 2. The proportion of Tier 1 sub-volumes reassigned to Tier 2 continued to increase with increasing relative position on the Tier 1 segment. Due to the complexity of COPD, not all GOLD 2 subjects will demonstrate the same trends over the 5-year period. As such, we evaluated an additional participant diagnosed with GOLD 2 COPD at baseline and at the 5-year follow-up visit. Details for this participant are provided in **Supplemental Fig. 1.** Despite having the same GOLD stage as the prior participant, this subject had significantly higher percent volume of PRM^Emph^ at baseline (11%), which increased by 6% (17%) at follow-up. As with the prior participant, we observed tier advancement at Tiers 0 through 2. In contrast, a much larger proportion of local lung scored Tier 1 (21%) advanced to Tier 3. Evaluating ePRM within individual subjects shows Tier 1 acts not only as an early indicator of disease but also as a key bridge between normal and obstructive parenchyma.

**Fig. 2:**
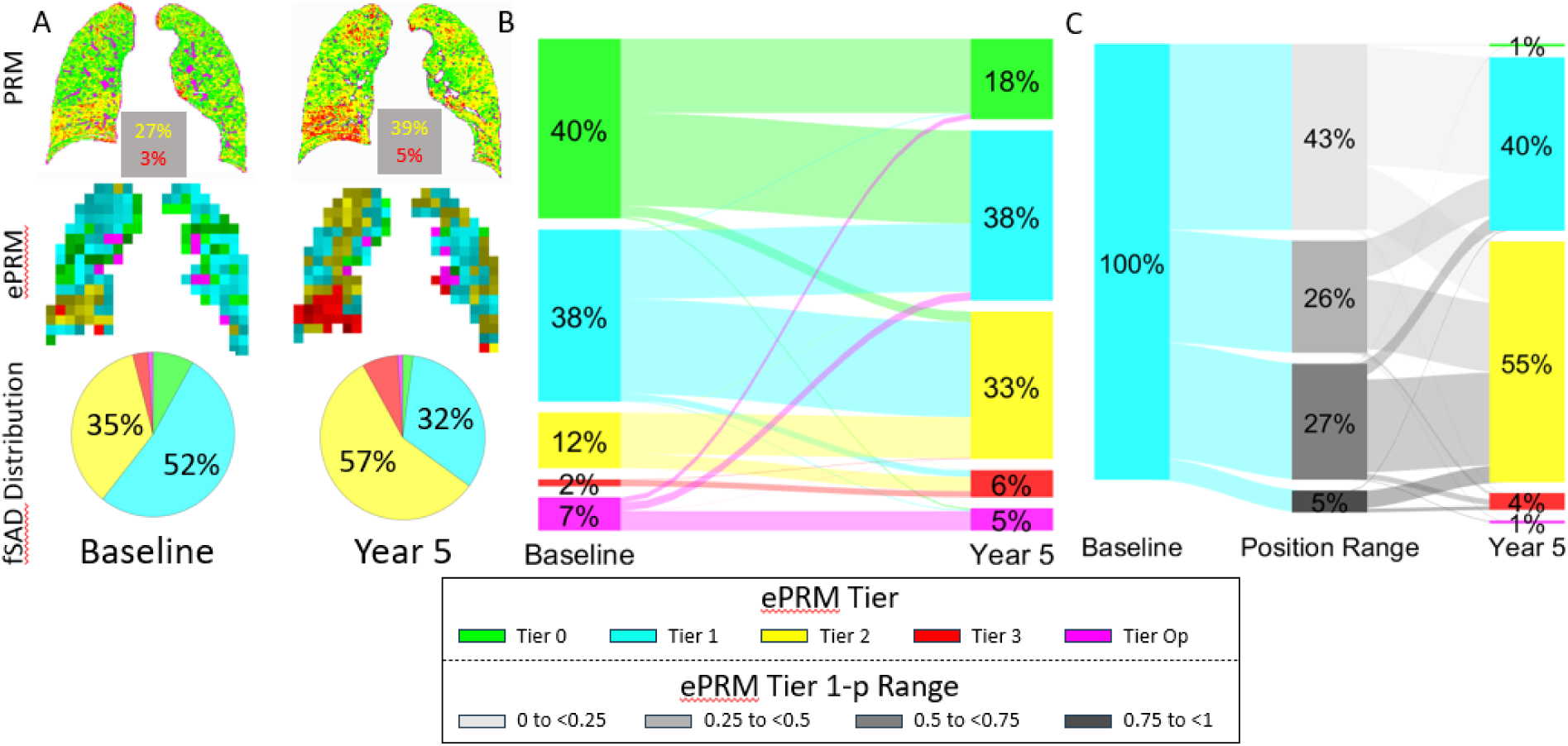
Longitudinal assessment of ePRM in representative subject with GOLD 2 COPD. (a) Representative coronal slices of serial PRM, with percent volume of fSAD (yellow) and emphysema (red) provided, and ePRM are presented from a participant diagnosed with GOLD 2 and 3 COPD at baseline and 5-year follow-up, respectively. The pie charts indicate the relative contribution of fSAD within each tier at the two time points. (b) Sub-volume transition between tiers over the 5-year period is depicted in a Sankey plot. The percent volume of each tier is shown at baseline and follow-up, with the flow lines indicating the direction and quantity of flow. The width of each line is proportional to the amount of transitioning sub-volumes, with thicker lines indicating larger quantities. (c) Transition of Tier 1 sub-volumes based on tier position (Tier 1-p) are presented in a Sankey plot. Here, sub-volumes are stratified into quarters based on their relative position in Tier 1. Subject 1 had a FEV1pp and FEV1/FVC of 51% and 0.5, respectively, at baseline and 41% and 0.41, respectively, at year 5.

### Local lung advances along a tier score towards obstructive tiers

We evaluated whether local lung with an unchanged tier score will demonstrate changes in tier position that bias progression towards Tiers 2 and 3. We first evaluated changes in tier mean position of regional lung retained on a single tier over 5 years (no reassignment). Irrespective of COPD severity or tier assignment, we observed a significant shift in mean position towards higher tier scores (**Fig. 3**), as indicated by the x-intercept deviating from the midpoint (0.5) in our waterfall plots. Differences between mean position values at years 0 and 5 were significant (**Supplemental Table 2**). Nevertheless, the magnitude of the mean position difference over 5 years varied between COPD subgroups in each tier. For local lung scored Tier 0 or 1, the No COPD subgroup had a significantly smaller change in tier position compared to the COPD subgroups (**Supplemental Table 2**). In contrast, for local lung scored Tier 2, mean change in tier position was significantly higher for No COPD and GOLD 1 and 2 compared to GOLD 3 and 4. Only local lung scored Tier 3 demonstrated significant differences in mean position across all subgroups, where higher COPD severity was associated with a significant increase in mean position (0.02±0.16 for No COPD, 0.08±0.17 for GOLD 1 and 2, and 0.10±0.16 for GOLD 3 and 4). On average, local lung with an unchanged tier score demonstrated changes in mean position towards Tiers 2 and 3.

**Fig. 3:**
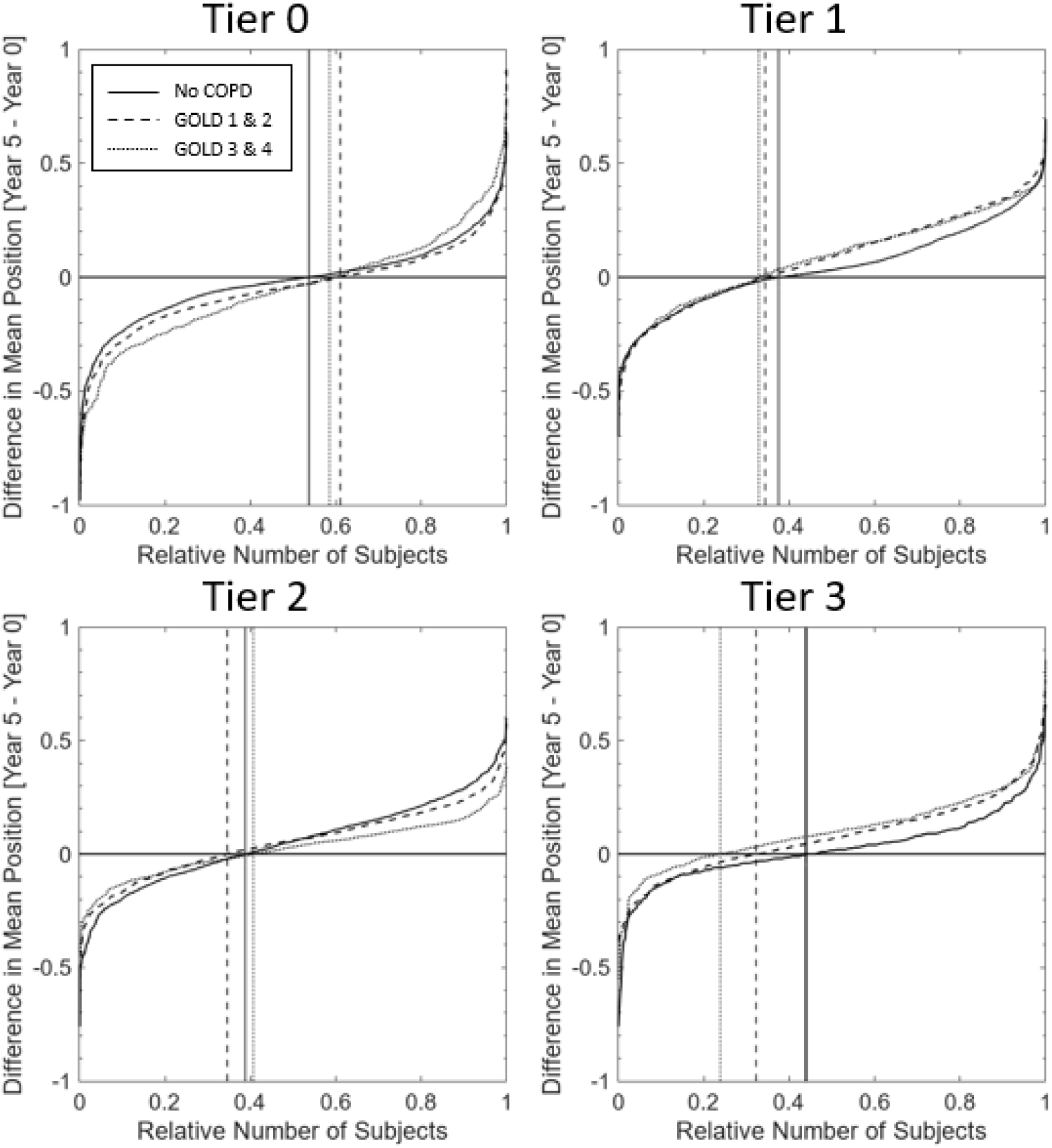
Longitudinal analyses of tier position. Waterfall plots illustrate the changes in whole-lung mean relative position for sub-volumes that remain in the same tier over a 5-year period. Participants were separated by GOLD subgroups: No COPD (PRISm and “at-risk”), GOLD 1 and 2, and GOLD 3 and 4. X-axis is the relative number of each subject. Four panels are provided for Tiers 0 through 3. Values in the x-intercept > or < 0.5 indicate a preference in movement direction along tier. Vertical lines indicate x-intercept equal to zero.

### Tier 1 position is a key indicator of advancement to obstructive tiers

We sought to determine whether the baseline mean Tier 1 position of local lung predicts reassignment to Tiers 2 or 3 at 5-year follow-up for all GOLD subgroups (**Fig. 4**). Using Receiver Operating Characteristic analysis, we found that the area under the curve (AUC) increased with increasing COPD severity (0.86 [sensitivity = 0.79, specificity = 0.80, optimal mean position cutoff = 0.22] for No COPD; 0.90 [0.80, 0.86, 0.39] for GOLD 1-2; and 0.92 [0.82, 0.83, 0.47] for GOLD 3-4). In contrast, local lung position on Tiers 0 and 2 did not predict advancement (**Fig. 4**), with AUC values not exceeding 0.6 for any subgroups. These findings suggest that local lung that advances along Tier 1 has an increased probability of advancing to obstructive tiers.

**Fig. 4:**
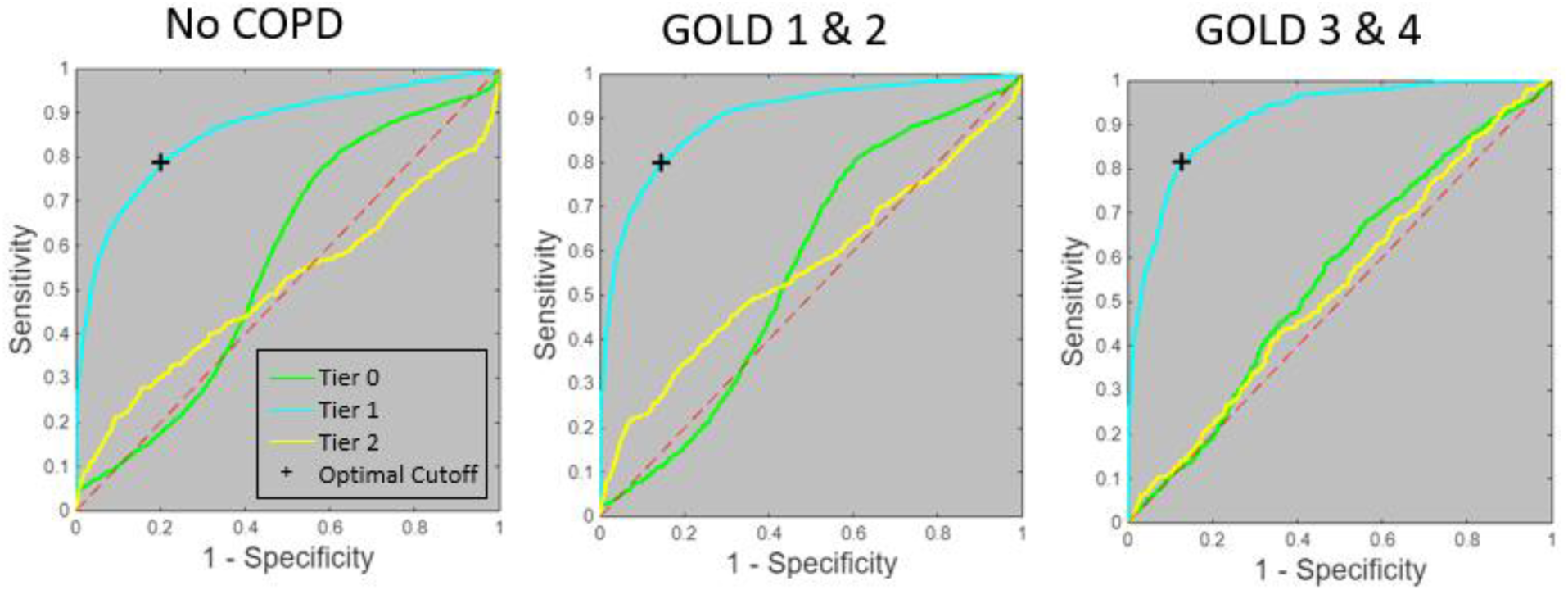
ROC analysis of Tier 1 position to predict tier score advancement. Receiver operator characteristic plot shows the potential of mean position at baseline for a given tier to predict the assignment of a sub-volume to a progressive tier at year 5. Outcome variable was defined as sub-volumes that progress and those that were remissive over five years. As an example, sub-volumes at Tier 1 at baseline were defined as 1 if reassigned to Tiers 2 and 3 at year 5. The remaining sub-volumes, those that remained in Tier 1 or withdrew to Tiers 0 or Op, were defined as 0. Analysis was performed separately for each COPD severity subgroup.

### Transition of local lung to obstructive Tiers increases with increasing COPD severity

We wanted to know if GOLD status affects the transition of regional lung between tier scores. As seen in our Sankey plots (**Fig. 5**), lung regions of individuals without COPD were primarily composed of Tier 0 (mean ± standard error of the mean: 62.8±0.49%) followed by Tier Op (24.8±0.51%) and Tier 1 (10±0.23%), which slightly changed over 5 years. A sizeable portion of Tier 0 lungs (9.3±0.28%) advanced to Tier 1, whereas lung regions scored Tier 1 were twice as likely to revert to Tier 0 (24.9±0.55%) than advance to Tier 2 (11.8±0.44%). Compared to participants without COPD, those with GOLD 1-2 COPD showed a higher prevalence of local lung advancing to higher tiers. On average, 23±0.59% of Tier 0 progressed to Tier 1, whereas 17.6±0.64% of Tier 1 improved to Tier 0 and 19.1±0.64% advanced to Tier 2. We observed a clear trend towards tier advancement over 5 years in participants with GOLD 3-4 COPD. At baseline, GOLD 3-4 participants had lungs consisting primarily of Tiers 1 (28.1±0.6%), 2 (21.5±0.74%) and 3 (24.9±1.14%). Nearly a quarter of Tiers 1 and 2 advanced to Tiers 2 (23.1±0.92%) and 3 (22.3±0.99%), respectively. The percent volumes of tiers at year 0 and year 5 and percent reassignments are provided in **Supplemental Table 3**. It appears that local lung reassigned to an advanced tier score is associated with a decline in lung function.

**Fig. 5:**
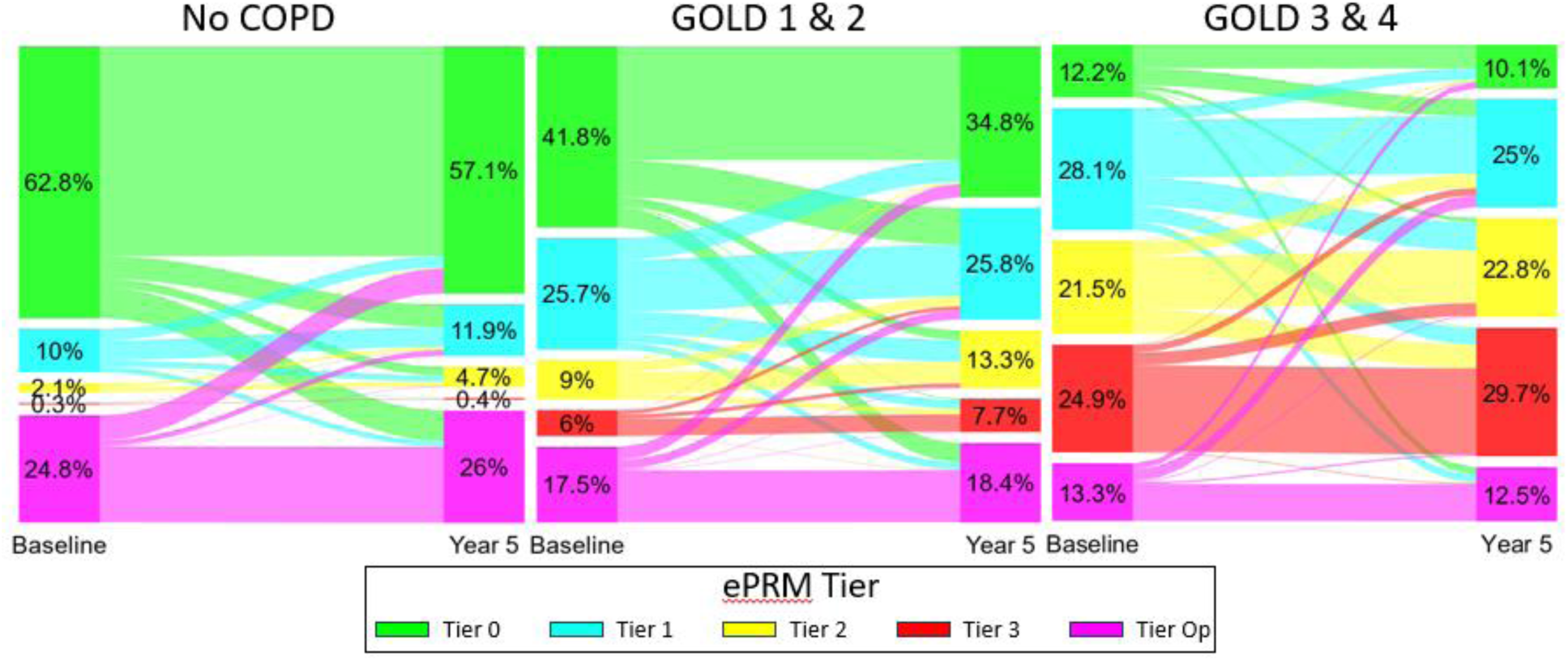
Evolution of sub-volume tier assignment by COPD severity. Sankey plots are shown for subgroups No COPD (PRISm and “at-risk”), GOLD 1 and 2, and GOLD 3 and 4. The mean percent volume is provided for each tier at each time point, where flow lines indicate the transition of sub-volumes between tier assignments from year 0 to year 5. The width of each line is proportional to the amount of transitioning sub-volumes, with thicker lines indicating larger quantities. To fit in the Sankey plots, real flow line values were normalized to year 0 and year 5 relative volumes for each tier. Means and standard error of the means of the tier percent volumes and actual transition values are provided in **Supplemental Table 3**.

### Retention of Tier 1 lung over 5 years was associated with improved lung function

We evaluated the relationship between baseline percent volume of local lung tier score and reassignment to new tiers to changes in FEV_1_ over 5 years. We analyzed each tier and subgroup separately. Only Tiers 0 and 1 had significant predictors for subjects without COPD (**Table 2**). Baseline percent volume of Tier 1 was negatively associated with lung function, such that a 1% increase in the percent volume of Tier 1 resulted in a 2.44 mL (p<0.001) drop in FEV_1_. In contrast, FEV_1_ improved with increases in the percent volume of lung that retained a Tier 1 score over 5 years. In the COPD subgroups, the percent volume of Tiers 2 and 3 at baseline or reassigned to these tiers were found to negatively impact FEV_1_. The only predictors that were significant for all subgroups were observed in Tiers 0 and 1. Although local lung scored Tier 1 is associated with early onset of PRM^fSAD^, the retention or reversal of Tier 1 score had a positive impact on lung function regardless of GOLD status.

**Table 2:** Evaluate tier transition to change in FEV_1_ (mL).

| A. No COPD |  | Percent Volume of Reassignment at Year 5 |  |  |  |  |
| --- | --- | --- | --- | --- | --- | --- |
|  |  | Percent Volume | Tier 0 | Tier 1 | Tier 2 | Tier 3 |
| Assignment at Year 0 | Tier 0 | 0.005 (0.986) | **2.926 (<0.0001)** | 1.077 (0.041) | **3.582 (<0.0001)** | -28.529 (0.146) |
|  | Tier 1 | **-2.435 (<0.0001)** | **1.194 (0.001)** | **1.311 (<0.0001)** | **1.104 (0.005)** | 2.164 (0.045) |
|  | Tier 2 | -2.61 (0.021) | -0.037 (0.908) | -0.342 (0.072) | 0.04 (0.813) | 1.57 (0.01) |
|  | Tier 3 | -6.782 (0.085) | -0.57 (0.314) | -0.272 (0.173) | -0.666 (0.026) | 0.329 (0.062) |
| B. GOLD 1 & 2 |  | Percent Volume of Reassignment at Year 5 |  |  |  |  |
|  |  | Percent Volume | Tier 0 | Tier 1 | Tier 2 | Tier 3 |
| Assignment at Year 0 | Tier 0 | **2.932 (<0.0001)** | **2.82 (<0.0001)** | -1.067 (0.114) | 1.827 (0.034) | -9.804 (0.039) |
|  | Tier 1 | **-5.093 (<0.0001)** | **3.924 (0.001)** | **3.55 (0.001)** | 2.214 (0.028) | -1.775 (0.163) |
|  | Tier 2 | **-4.728 (<0.0001)** | -0.737 (0.393) | -0.001 (0.998) | -0.462 (0.222) | **-3.268 (<0.0001)** |
|  | Tier 3 | **-5.508 (<0.0001)** | -1.378 (0.377) | -0.122 (0.758) | -0.973 (0.017) | **-0.966 (<0.0001)** |
| C. GOLD 3 & 4 |  | Percent Volume of Reassignment at Year 5 |  |  |  |  |
|  |  | Percent Volume | Tier 0 | Tier 1 | Tier 2 | Tier 3 |
| Assignment at Year 0 | Tier 0 | **3.7 (<0.0001)** | **2.528 (<0.0001)** | -0.64 (0.286) | -0.267 (0.778) | -3.011 (0.208) |
|  | Tier 1 | -1.032 (0.258) | **10.014 (<0.0001)** | **3.725 (0.007)** | 3.272 (0.012) | -0.048 (0.973) |
|  | Tier 2 | -0.84 (0.265) | **8.691 (<0.0001)** | **-3.739 (0.003)** | **-3.191 (0.004)** | **-5.003 (<0.0001)** |
|  | Tier 3 | **-2.192 (0.001)** | 0.835 (0.704) | 0.174 (0.844) | -1.159 (0.087) | **-1.984 (<0.0001)** |

**Note:** FEV<sub>1</sub> is change in mL and ePRM readouts are in percent volume.
Values are parameter estimates (P values) for relative volume of tiers at baseline and relative value of transition between tiers evaluated in separate linear mixed effects models (rows). ePRM parameters are in the range [0 100%]. Linear mixed effects models were performed using data from the subgroups: A. No COPD, B. GOLD 1 and 2, and C. GOLD 3 and 4. The dependent variable was the change in FEV<sub>1</sub> (mL) over 5-year interval [FEV<sub>1</sub>(5 years) – FEV<sub>1</sub>(0 years)]. Predictors included the ePRM parameters in the header and baseline age, sex, BMI, and FEV<sub>1</sub>, as well as change in ATS smoking pack-years, smoking status, and scanner make over 5-year interval. The subject ID was used as the random effect. Bold values indicate statistical significance accounting for multiple parameters (0.05/5 = 0.01).

## Discussion

In this large prospective analysis of individuals with COPD or at risk for developing COPD, we demonstrate that chest CT ePRM can categorize local lung tissue into distinct tiers of disease severity that reflect reversible and irreversible processes and correlate with subsequent changes in pulmonary function. ePRM translates quantitative CT readouts into a disease severity scale while retaining spatial information, providing a comprehensive tool for clinicians to assess and monitor disease progression. The ability to radiographically identify and quantify lung tissue by risk category could not only help clinicians identify the most effective regimens of currently available treatments for patients but also open up the potential for locally targeted therapeutics.

Notably, we identified Tier 1 as a critical two-way bridge between healthy and diseased lung tissue (**Fig. 1**). Although COPD generally progresses over time, our data suggest this is due not to uniform irreversible damage, but to an imbalance between lung regions that progress to more advanced tiers of disease and regions that revert to less obstructive tiers. For example, among individuals with mild and moderate COPD (GOLD 1-2), we show that 25.0% of sub-volumes in Tier 1 progress to Tiers 2 and 3, whereas 17.6% revert to Tier 0 (**Fig. 5** and **Supplemental Table 3**). For those who revert to Tier 0, existing therapies can reduce airway inflammation and improve FEV_1_, which may be what we are detecting at a local level. What is crucial to understand are the longer-term outcomes of these reversible regions and the ability of therapies to meaningfully modify the natural trajectory of the disease [26]. Such therapies are desperately needed, since COPD remains a leading cause of morbidity and mortality worldwide [1–3]. Moreover, unlike other leading causes of death such as cancer and cardiovascular disease that now show improved prognoses, COPD mortality rates have not declined over the past two decades [27, 28]. Therefore, understanding the intersection of genetic, environmental (e.g., cigarette smoking pattern [29], occupational exposures [30], air pollution [31]), structural (e.g., lung dysynapsis [32], mucus plugging [33]), physiologic (e.g., local inflammatory milieu and immune regulation [17]) and therapeutic determinants of changes in local lung composition, as detected by ePRM, will have significant implications for COPD care. We observed that a higher proportion of sub-volumes either transitioning from Tier 1 to Tier 0 or remaining in Tier 1 over five years was associated with reduced FEV_1_ decline—a finding that underscores the role of Tier 1 as an important bridge in the natural progression of COPD. This trend persisted across all disease categories, including participants with GOLD 1-2 and GOLD 3-4 COPD and those at risk for developing COPD, even after adjusting for important confounders such as smoking history and baseline FEV_1_ (**Table 2**). Additionally, we demonstrated the importance of the position of sub-volumes on the Tier 1 segment. Sub-volumes on all tiers tended to migrate towards more obstructive tiers over 5 years (**Fig. 3**). Regardless of GOLD status, local lung with a mean Tier 1 position greater than 0.5 were found to progress to less reversible Tiers 2 and 3 (**Fig. 4**). In contrast, mean sub-volume positions on Tiers 0 and 2 lacked similar discriminative accuracy. These findings suggest that local lung scored as Tier 1 are most susceptible to disease progression and represent a pool of lung that should be protected with effective behavioral and therapeutic interventions at any stage of the disease. Whether the early initiation of novel biologic therapies targeting inflammation can help preserve this potentially reversible pool of lung in the long term remains to be determined [34–36]. In this setting, ePRM may be a powerful application for identifying which at-risk individuals are most likely to develop COPD, which is currently challenging to estimate with available clinical tools [37, 38].

We acknowledge limitations to our study. First, we measured changes in lung sub-volume composition only at baseline and 5 years later, the timepoints when scans were obtained. How this composition changed over shorter intervals, such as yearly, during the 5-year period remains unknown. Second, we did not account for pharmacologic and non-pharmacologic therapies participants received during the study. Estimating the impact of therapies on our findings is challenging, as each participant may have received varying combinations of treatments over the follow-up period. Third, our elastic principal graph model relies on the PRM composition of lung sub-volumes. Future models may incorporate additional CT-derived quantitative metrics, such as blood perfusion or airway wall thickness, to potentially enhance their predictive capability [39–41]. Fourth, while our outcome of interest, FEV_1_ change, served as a solid initial benchmark for testing our model, the value of ePRM should be explored for other important COPD-related outcomes, including the frequency of respiratory exacerbations, the progression of respiratory symptom burden, and long-term mortality [42].

To summarize, ePRM is an innovative quantitative CT scoring technique for precisely and reliably monitoring local COPD progression. This imaging biomarker provides a method for detecting and characterizing COPD earlier in its course, when altering the natural trajectory of the disease may still be possible [43]. The proportion and mean position of sub-volumes in Tier 1 play a particularly important role in this context. Leveraging ePRM for early COPD detection and clinical trial enrichment has the potential to shift the paradigm from the current status quo toward disease interception and modification [44].

## Methods

### Subject and clinical data

Demographic, clinical and CT data were collected on subjects who participated in both Phase 1 (baseline) and Phase 2 (approximately 5 years later) of the COPDGene study. The COPDGene trial enrolled individuals 45-80 years of age with ≥ 10 pack-years of current or former tobacco use [9]. Of the 4,585 Phase 2 participants, a total of 3,631 had adequate CT acquisitions and clinical data at both examination sessions for our study. A list of clinical metrics and the methods for acquiring, aligning, and processing inspiratory and expiratory CT scans to generate PRM data are provided in the **Supplemental Methods**. Institutional review boards of COPDGene study sites approved the study protocol, and all participants gave written informed consent.

### Elastic PRM (ePRM)

PRM data at both time points for all cases were processed using ePRM. The ePRM workflow, which classifies each lung parenchyma sub-volume into one of five tiers (**Fig. 1**), consists of three steps. (*Step 1*) All PRM were resampled using a nearest neighbor method to a matrix size of 512^3^ and separated into non-overlapping sub-volumes of 16^3^ voxels per sub-volume (∼1 µL). All sub-volumes with less than 75% lung were omitted. The percent volumes for all PRM classifications (PRM^Norm^, PRM^fSAD^, PRM^Emph^ and PRM^PD^) were calculated in each sub-volume, then organized in tabular form. (*Step 2*) Using dedicated ElPiGraph functions [23], we projected each sub-volume to the elastic principal graph model to determine its a) tier assignment and b) relative position within its tier. Tier assignment consists of five categories: Tiers 0 through 3 and Op (Op indicates opacities associated with high attenuation areas). Tier relative position indicates the location of a given sub-volume along a tier. All terminal branches (Tiers 0, 2, 3, and Op as shown in **Fig. 1**) are designated 0 at their junction with Tier 1 and 1 at their terminus. Tier 1, the bridge, is designated 0 at its junction with Tiers 0 and Op and 1 at its junction with Tiers 2 and 3. (*Step 3*) Each sub-volume was mapped onto the tree and resampled back to the original 3D PRM space (**Fig. 1**). We use the shorthand “Tier ***Assignment***-p” to indicate position along a given tier (e.g., “Tier 1-p” indicates relative position of a sub-volume along Tier 1). Whole lung measurements for each participant are provided as: 1) the percent volume of each tier assignment calculated as 100 times the sum of tier-like sub-volumes over all sub-volumes and 2) the mean relative position across all like-tiers in the lungs.

### Building ePRM model

The model for our ePRM tiered system was built using the ElPiGraph machine learning algorithm. Details of this method have been described [23]. In brief, we developed our ePRM model using 300 randomly selected participants from Phase 1 COPDGene with equal sex representation (male/female = 150/150). As PRM^Norm^ is overrepresented in participants without COPD or with mild disease, we selected cases per Global Initiative for Chronic Obstructive Lung Disease (GOLD) spirometry grade with a weight based on 100% minus the percent volume of PRM^Norm^ within the lungs [= 100% − %PRM^Norm^]. Participant numbers and characteristics per GOLD status are provided in **Supplemental Table 1**. From each of the 300 cases, we randomly selected 10,000 sub-volumes (N = 300 × 10,000 = 3,000,000 sub-volumes), allowing for overlap but non-repeating. Sub-volumes with a percent volume of lung < 75% were omitted, leaving 2,750,000 sub-volumes. The percent volume of PRM classes were calculated for each sub-volume and organized in tabular form (sub-volumes along rows and PRM percent volumes along columns). A final model was determined by processing the tabulated data using ElPiGraph with the following parameters: three principal components based on the Broken-Stick model, 100 fits, 10% of data randomly selected for each fit (N = 275,000 sub-volumes per run), and 40 nodes based on analysis of the balance between the percent of data variance explained by model versus complexity of model.

### Data and statistical analyses

All continuous variables were presented as the mean ± the standard error of the mean. Categorical variables were presented as counts and percentages. Unless stated otherwise, all analyses were performed on three subgroups of participants based on GOLD spirometry grade [45]: No COPD (no airflow obstruction, either “at-risk” or PRISm, Preserved Ratio Impaired Spirometry [46]), GOLD 1-2 (mild and moderate airflow obstruction), and GOLD 3-4 (severe and very severe airflow obstruction). All data and statistical analyses, as well as pre- and post-processing of model projection of data, were performed in MATLAB version 2024a (MathWorks, Natick, MA, USA). Computing the elastic principal graph and projection of tabulated sub-volume data from all subjects was performed using ElPiGraph Python package installed from https://github.com/j-bac/elpigraph-python.

#### Assessment of tier position change over time

We assessed how the relative position of sub-volumes changed from years 0 and 5 in three GOLD groups of COPD severity. Only sub-volumes that remained in their original tier across the two time points were evaluated. For each participant, we calculated the difference in the mean relative positions of all sub-volumes within each tier by comparing their values at baseline and year 5 (example for a single participant: = Mean Relative Position of sub-volumes in Tier 1 in the lungs at year 5 − Mean Relative Position of sub-volumes in Tier 1 in the lungs at year 0). The difference between mean relative positions at each time point was assessed using a paired Student’s t-test. We also assessed whether the difference in mean position over time varied between COPD subgroups. This analysis was performed using a Kruskal-Wallis test adjusted for multiple comparisons. Both statistical tests were assumed significant at p<0.05.

#### Association between baseline mean relative position and subsequent tier advancement

This analysis was performed for sub-volumes originating in Tiers 0 through 2 at baseline. Advancement (binary outcome of 0 or 1) was defined as reassignment to a higher tier at year 5; Tier Op was considered equivalent or prior to Tier 0. As an example, sub-volumes that originate on Tier 0 would be labeled as “1” if they advance to Tiers 1 through 3 at year 5 or “0” if they remain on Tier 0 or remiss to Tier Op. In MATLAB, a logistic regression model was used with the baseline mean relative position on a tier as the sole predictor. Receiver operating characteristic analysis was used to determine the area under the curve, sensitivity, specificity and optimal cutoff, as defined at the maximum Youden’s Index (J = Sensitivity + Specificity −1), for the regression results for Tiers 0, 1 and 2. As before, we stratified analyses by COPD GOLD subgroups. In addition, we determined the probability of tier score advancement for different cutoffs of local lung Tier 1 position. The range of cutoff values were from the optimal cutoff determined from the ROC analysis to 1, with intervals of 0.1. This analysis was performed by applying the logistic regression model to local lung Tier 1 position values greater than or equal to a specific cutoff.

#### Longitudinal assessment of ePRM tier assignment

A single 3D sub-volume consists of a tier assignment and a relative position at two time points, years 0 and 5. As there are five tiers in our ePRM model, there are 25 possible combinations of reassignments over the 5-year period. For example, a sub-volume in Tier 1 at baseline may be reassigned to Tiers 0, 2, 3 or Op, or remain in Tier 1 (5 possibilities) at year 5. We calculated the percent volume of reassignment for a given tier at year 0 as the number of sub-volumes reassigned to any tier at year 5, normalized by the total number of sub-volumes within that tier at year 0. We used Sankey plots to illustrate the reassignment of tiers between years 0 and 5. As the data presented are averaged over multiple subjects within one of the three GOLD subgroups, the flow lines on the Sankey plots were determined by normalizing the row and columns of the transition matrix (i.e., percent volume of reassignment) by the percent volume of tiers at years 0 and 5, respectively.

#### Evaluate the effects of percent volume of baseline tier and reassignment on FEV_1_ change

To test this association, we used linear mixed effects models with the outcome defined as the change in FEV_1_ over 5 years [FEV_1_(year 5) – FEV_1_(year 0)]. Our analysis was performed separately for each tier (Tiers 0 through 3) and GOLD subgroup (no COPD, GOLD 1-2 and GOLD 3-4). Predictors of interest were percent volume of the tier at baseline (e.g. percent volume of Tier 1 in the lungs) and percent volume of local lung reassigned to a tier at follow-up (e.g. percent volume of lung that transitions over 5 years: Tier 1→Tier 0, Tier 1→Tier 1, Tier 1→Tier 2, and Tier 1→Tier 3). We adjusted the models for age, sex, BMI, and FEV_1_ at baseline and change in smoking pack-years (year 5 – year 0), smoking status (0 for former smoker no change; 1 for former to current smoker; 2 for current to former; and 3 for current smoker no change) and scanner make (0 for same make and 1 for different make) over the time interval. Subject ID was used as a random intercept. We applied the Bonferroni correction to account for multiple comparisons, adjusting the p-value threshold to 0.01 (0.05/5 predictors of interest).

## Data availability

The datasets presented in this study are not readily available because they are part of NIH sponsored clinical trials and require signing of a data use agreement. For instructions on how to access COPDGene data visit https://www.copdgene.org/phase-1-study-documents.htm.

## Acknowledgments

This work was supported by the National Heart, Lung, and Blood Institute (NHLBI) of the National Institutes of Health (NIH) grants U01 HL089897 and U01 HL089856 and by NIH contract 75N92023D00011. The COPDGene study (<u>NCT00608764</u>) has also been supported by the COPD Foundation through contributions made to an Industry Advisory Committee that has included AstraZeneca, Bayer Pharmaceuticals, Boehringer-Ingelheim, Genentech, GlaxoSmithKline, Novartis, Pfizer and Sunovion. The authors would like to thank Lee Olsen for editing the manuscript.

## Author contributions

MKH, SG and CJG conceived the study and hypotheses. WWL and CJG managed data preparation, statistical analysis, software implementation, experimentation, and presentation of results. WWL and CJG wrote the initial draft of the manuscript. MKH and CJG supervised the project. MKH and WWL, FJM, and EAK provided domain expert advice and evaluation of the project. SM provided consultancy on the statistical analyses. CRH processed COPDGene CT data for lung segmentation and image registration. AZ, EMM, and ANG supported algorithmic development and implementation of elastic principal graphs for this project. AZ provided expert support in the theory and application of elastic principal graphs and related data analysis. All authors provided substantive, critical reviews and approved of the submitted manuscript.

## References

1. Collaborators, G.B.D.C.o.D., Global burden of 288 causes of death and life expectancy decomposition in 204 countries and territories and 811 subnational locations, 1990-2021: a systematic analysis for the Global Burden of Disease Study 2021. Lancet, 2024. 403(10440): p. 2100–2132.

2. Collaborators, G.B.D.C.R.D., Prevalence and attributable health burden of chronic respiratory diseases, 1990-2017: a systematic analysis for the Global Burden of Disease Study 2017. Lancet Respir Med, 2020. 8(6): p. 585–596.

3. Boers, E., et al., Global Burden of Chronic Obstructive Pulmonary Disease Through 2050. JAMA Netw Open, 2023. 6(12): p. e2346598.

4. Duffy, S., M. Weir, and G.J. Criner, The complex challenge of chronic obstructive pulmonary disease. Lancet Respir Med, 2015. 3(12): p. 917–9.

5. Labaki, W.W., et al., The Role of Chest Computed Tomography in the Evaluation and Management of the Patient with Chronic Obstructive Pulmonary Disease. Am J Respir Crit Care Med, 2017. 196(11): p. 1372–1379.

6. Bhatt, S.P., et al., Imaging Advances in Chronic Obstructive Pulmonary Disease. Insights from the Genetic Epidemiology of Chronic Obstructive Pulmonary Disease (COPDGene) Study. Am J Respir Crit Care Med, 2019. 199(3): p. 286–301.

7. Galbán, C.J., et al., Computed tomography-based biomarker provides unique signature for diagnosis of COPD phenotypes and disease progression. Nat Med, 2012. 18(11): p. 1711–5.

8. Vasilescu, D.M., et al., Noninvasive Imaging Biomarker Identifies Small Airway Damage in Severe Chronic Obstructive Pulmonary Disease. Am J Respir Crit Care Med, 2019. 200(5): p. 575–581.

9. Regan, E.A., et al., Genetic epidemiology of COPD (COPDGene) study design. COPD, 2010. 7(1): p. 32–43.

10. Couper, D., et al., Design of the Subpopulations and Intermediate Outcomes in COPD Study (SPIROMICS). Thorax, 2014. 69(5): p. 491–4.

11. Bell, A.J., et al., Local heterogeneity of normal lung parenchyma and small airways disease are associated with COPD severity and progression. Respir Res, 2024. 25(1): p. 106.

12. Hogg, J.C., P.T. Macklem, and W.M. Thurlbeck, Site and nature of airway obstruction in chronic obstructive lung disease. N Engl J Med, 1968. 278(25): p. 1355–60.

13. McDonough, J.E., et al., Small-airway obstruction and emphysema in chronic obstructive pulmonary disease. N Engl J Med, 2011. 365(17): p. 1567–75.

14. Bhatt, S.P., et al., Association Between Functional Small Airways Disease and FEV Decline in COPD. Am J Respir Crit Care Med, 2016.

15. Labaki, W.W., et al., Voxel-Wise Longitudinal Parametric Response Mapping Analysis of Chest Computed Tomography in Smokers. Acad Radiol, 2019. 26(2): p. 217–223.

16. Boes, J.L., et al., Parametric response mapping monitors temporal changes on lung CT scans in the subpopulations and intermediate outcome measures in COPD Study (SPIROMICS). Acad Radiol, 2015. 22(2): p. 186–94.

17. Hogg, J.C., et al., The nature of small-airway obstruction in chronic obstructive pulmonary disease. N Engl J Med, 2004. 350(26): p. 2645–53.

18. Hogg, J.C., P.D. Pare, and T.L. Hackett, The Contribution of Small Airway Obstruction to the Pathogenesis of Chronic Obstructive Pulmonary Disease. Physiol Rev, 2017. 97(2): p. 529–552.

19. Zhou, Y., et al., Tiotropium in Early-Stage Chronic Obstructive Pulmonary Disease. N Engl J Med, 2017. 377(10): p. 923–935.

20. Albergante, L., et al., Robust and Scalable Learning of Complex Intrinsic Dataset Geometry via ElPiGraph. Entropy (Basel), 2020. 22(3).

21. Gorban, A.N., N.R. Sumner, and A.Y. Zinovyev, Topological grammars for data approximation. Applied Mathematics Letters, 2007. 20(4): p. 382–386.

22. Zinovyev, A. and E. Mirkes, Data complexity measured by principal graphs. Computers & Mathematics with Applications, 2013. 65(10): p. 1471–1482.

23. Golovenkin, S.E., et al., Trajectories, bifurcations, and pseudo-time in large clinical datasets: applications to myocardial infarction and diabetes data. Gigascience, 2020. 9(11).

24. Chen, H., et al., Single-cell trajectories reconstruction, exploration and mapping of omics data with STREAM. Nat Commun, 2019. 10(1): p. 1903.

25. Bell, A.J., et al., Temporal Exploration of COPD Phenotypes: Insights from the COPDGene and SPIROMICS Cohorts. Am J Respir Crit Care Med, 2024.

26. Agusti, A. and J.C. Hogg, Update on the Pathogenesis of Chronic Obstructive Pulmonary Disease. N Engl J Med, 2019. 381(13): p. 1248–1256.

27. Ma, J., et al., Temporal Trends in Mortality in the United States, 1969-2013. JAMA, 2015. 314(16): p. 1731–9.

28. Collaborators, G.U.H.D., Cause-specific mortality by county, race, and ethnicity in the USA, 2000-19: a systematic analysis of health disparities. Lancet, 2023. 402(10407): p. 1065–1082.

29. Oelsner, E.C., et al., Lung function decline in former smokers and low-intensity current smokers: a secondary data analysis of the NHLBI Pooled Cohorts Study. Lancet Respir Med, 2020. 8(1): p. 34–44.

30. Paulin, L.M., et al., Occupational Exposures and Computed Tomographic Imaging Characteristics in the SPIROMICS Cohort. Ann Am Thorac Soc, 2018. 15(12): p. 1411–1419.

31. Sin, D.D., et al., *Air pollution and COPD: GOLD* 2023 committee report. Eur Respir J, 2023. 61(5).

32. Smith, B.M., et al., Association of Dysanapsis With Chronic Obstructive Pulmonary Disease Among Older Adults. JAMA, 2020. 323(22): p. 2268–2280.

33. Diaz, A.A., et al., Airway-Occluding Mucus Plugs and Mortality in Patients With Chronic Obstructive Pulmonary Disease. JAMA, 2023. 329(21): p. 1832–1839.

34. Bhatt, S.P., et al., Dupilumab for COPD with Type 2 Inflammation Indicated by Eosinophil Counts. N Engl J Med, 2023. 389(3): p. 205–214.

35. Bhatt, S.P., et al., Dupilumab for COPD with Blood Eosinophil Evidence of Type 2 Inflammation. N Engl J Med, 2024. 390(24): p. 2274–2283.

36. Polverino, F. and D.D. Sin, Type 2 airway inflammation in COPD. Eur Respir J, 2024. 63(5).

37. Antuni, J.D. and P.J. Barnes, Evaluation of Individuals at Risk for COPD: Beyond the Scope of the Global Initiative for Chronic Obstructive Lung Disease. Chronic Obstr Pulm Dis, 2016. 3(3): p. 653–667.

38. Han, M.K., et al., From GOLD 0 to Pre-COPD. Am J Respir Crit Care Med, 2021. 203(4): p. 414–423.

39. Han, M.K., et al., Chronic obstructive pulmonary disease exacerbations in the COPDGene study: associated radiologic phenotypes. Radiology, 2011. 261(1): p. 274–82.

40. Johannessen, A., et al., Mortality by level of emphysema and airway wall thickness. Am J Respir Crit Care Med, 2013. 187(6): p. 602–8.

41. Pistenmaa, C.L., et al., Pulmonary Arterial Pruning and Longitudinal Change in Percent Emphysema and Lung Function: The Genetic Epidemiology of COPD Study. Chest, 2021. 160(2): p. 470–480.

42. Cazzola, M., et al., An Update on Outcomes for COPD Pharmacological Trials: A COPD Investigators Report - Reassessment of the 2008 American Thoracic Society/European Respiratory Society Statement on Outcomes for COPD Pharmacological Trials. Am J Respir Crit Care Med, 2023. 208(4): p. 374–394.

43. Labaki, W.W. and M.K. Han, Improving Detection of Early Chronic Obstructive Pulmonary Disease. Ann Am Thorac Soc, 2018. 15(Suppl 4): p. S243–S248.

44. Stolz, D., et al., Towards the elimination of chronic obstructive pulmonary disease: a Lancet Commission. Lancet, 2022. 400(10356): p. 921–972.

45. Agustí, A., et al., Global Initiative for Chronic Obstructive Lung Disease 2023 Report: GOLD Executive Summary. Eur Respir J, 2023. 61(4).

46. Wan, E.S., et al., Epidemiology, genetics, and subtyping of preserved ratio impaired spirometry (PRISm) in COPDGene. Respir Res, 2014. 15(1): p. 89.

