## Supplemental Figures, Tables, Methods for "Quantitative CT Scoring for Local COPD Severity"

### Supplemental Information Guide

| Item | Title/Description |
|---|---|
| Supplemental Table 1 | Randomly Selected Cases for Model Building |
| Supplemental Figure 1 | Additional Subject Diagnosed with GOLD 2 COPD |
| Supplemental Table 2 | Supporting data for Fig. 3 |
| Supplemental Table 3 | Supporting data for Fig. 5 |
| Supplemental Methods | Clinical Metrics, CT Parameters, Spatial Alignment of CT Scans and PRM |
| Supplemental Figure 2 | PRM Profiles along Elastic Principal Graph |

Supplemental Table 1

| | PRISm | 0 | 1 | 2 | 3 | 4 |
|---|---|---|---|---|---|---|
| N | 44 | 34 | 40 | 48 | 62 | 72 |
| Age Enrolled (yr) | 58 (4) | 58 (4) | 60 (4) | 60 (4) | 60 (4) | 61 (4) |
| Sex (m/f) | 22/22 | 17/17 | 20/20 | 24/24 | 31/31 | 36/36 |
| $PRM^{Norm}$ (%) | 54 (16) | 64 (13) | 60 (12) | 52 (11) | 34 (13) | 19 (10) |
| $PRM^{fSAD}$ (%) | 9 (7) | 9 (8) | 12 (8) | 20 (9) | 30 (12) | 36 (10) |
| $PRM^{Emph}$ (%) | 1 (1) | 1 (1) | 2 (3) | 4 (5) | 14 (12) | 28 (15) |
| $PRM^{PD}$ (%) | 36 (18) | 25 (13) | 24 (10) | 23 (10) | 20 (11) | 15 (6) |

Notes: Characteristics of the 300 participants randomly selected to build the Tiered Severity Scoring System. Continuous variables are presented as means (standard deviation). GOLD, Global Initiative for Chronic Obstructive Lung Disease; PRISm, preserved ratio impaired spirometry; and “at-risk”, at-risk smokers with normal spirometry. PRM values are presented as the percent volume of the lungs. To maximize cases with varying levels of $PRM^{Norm}$, $PRM^{fSAD}$, $PRM^{Emph}$ and $PRM^{PD}$, a weight was applied based on 100-percent volume of $PRM^{Norm}$.

#### Supplemental Figure 1

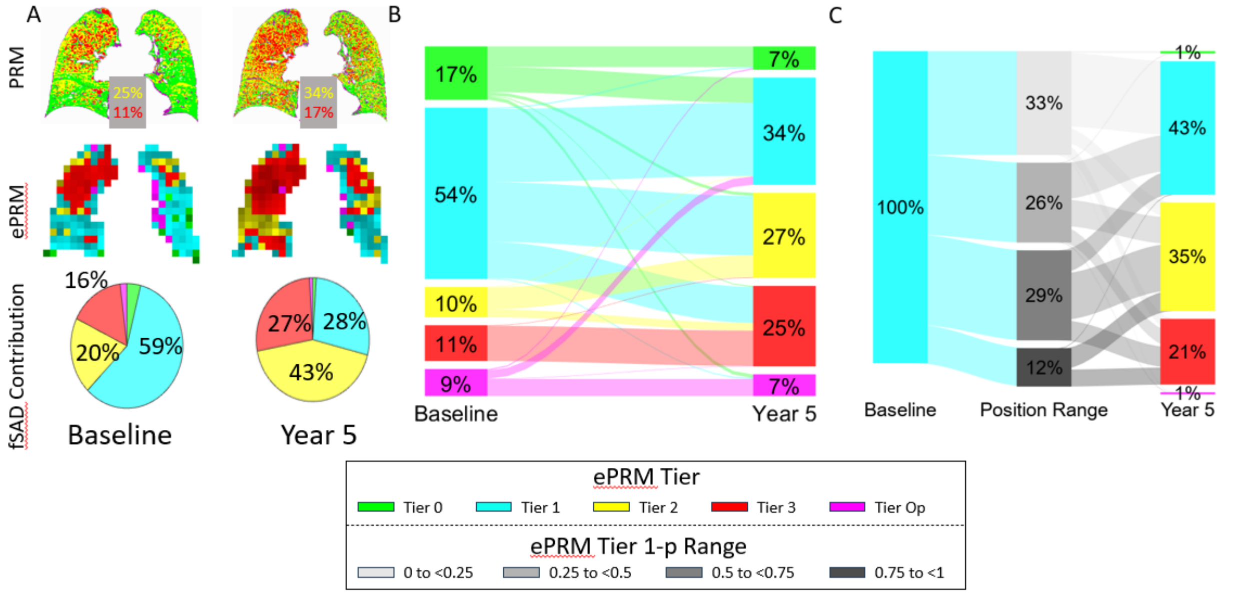


This subject, diagnosed with GOLD 2 COPD at baseline, had $FEV_1$ percent predicted of 57% and 53% at year 0 and 5, respectively, and an increase in $PRM^{fSAD}$ from 25% to 34% and $PRM^{Emph}$ from 11% to 17%. Regions of lung with $PRM^{Norm}$ (green voxels) progressed to $PRM^{fSAD}$ and $PRM^{Emph}$ (yellow and red voxels, respectively) across both lungs. Unlike the subject in **Fig. 2**, regions of Tier 1 appear to progress to both Tier 2 and 3. Of the 54% of Tier 1 lung, 35% evolved to Tier 2 and 21% to Tier 3.

#### Supplemental Table 2

| Tier | COPD | N | Mean Position Year 0 | Year 5 | Difference |
|---|---|---|---|---|---|
| 0 | No COPD | 2144 | 0.77 (0.13) | 0.76 (0.14)* | -0.02 (0.18) |
| | GOLD 1 & 2 | 1035 | 0.71 (0.15) | 0.66 (0.17)* | -0.05 (0.19)# |
| | GOLD 3 & 4 | 396 | 0.54 (0.18) | 0.49 (0.21)* | -0.05 (0.25)# |
| 1 | No COPD | 2110 | 0.29 (0.11) | 0.33 (0.12)* | 0.04 (0.18) |
| | GOLD 1 & 2 | 1046 | 0.32 (0.12) | 0.40 (0.13)* | 0.08 (0.21)# |
| | GOLD 3 & 4 | 424 | 0.41 (0.13) | 0.50 (0.12)* | 0.09 (0.20)# |
| 2 | No COPD | 1047 | 0.73 (0.12) | 0.78 (0.13)* | 0.05 (0.19) |
| | GOLD 1 & 2 | 908 | 0.74 (0.11) | 0.79 (0.12)* | 0.05 (0.16) |
| | GOLD 3 & 4 | 416 | 0.77 (0.09) | 0.79 (0.10)* | 0.02 (0.12)#,$ |
| 3 | No COPD | 530 | 0.41 (0.18) | 0.43 (0.17)* | 0.02 (0.16) |
| | GOLD 1 & 2 | 704 | 0.40 (0.16) | 0.47 (0.18)* | 0.08 (0.17)# |
| | GOLD 3 & 4 | 381 | 0.46 (0.17) | 0.56 (0.16)* | 0.10 (0.16)#,$ |

Note: Averages (standard deviation) are provided for the tier mean position and difference of the mean positions, separated by Tier and COPD subgroups. Significant differences between mean positions at years 0 and 5 were determined using paired t-test with * indicating significance at p<0.05. Significance of differences in mean position between COPD subgroups was determined using Kruskal-Wallis test with # and $ indicating significant difference from No COPD and GOLD 1 and 2, respectively, at p < 0.017 adjusted for multiple comparisons.

#### Supplemental Table 3

| | Percent Volume @ Year 0 | Reassignment at Year 5 | | | | | Percent Volume @ Year 5 |
|---|---|---|---|---|---|---|---|
| | | Tier 0 | Tier 1 | Tier 2 | Tier 3 | Tier Op | |
| | | | | No COPD | | | |
| Tier 0 | 62.8 (0.49) | 75.2 (0.52) | 9.3 (0.28) | 2.8 (0.22) | 0 (0.01) | 12.6 (0.4) | 57.1 (0.53) |
| Tier 1 | 10 (0.23) | 24.9 (0.55) | 48.2 (0.5) | 11.8 (0.44) | 1.1 (0.11) | 13.9 (0.36) | 11.9 (0.26) |
| Tier 2 | 2.1 (0.11) | 10.3 (0.47) | 37.3 (0.67) | 47.2 (0.79) | 3.1 (0.25) | 2.1 (0.2) | 4.7 (0.24) |
| Tier 3 | 0.3 (0.03) | 4.1 (0.33) | 32.4 (0.8) | 16.1 (0.59) | 42.6 (0.89) | 4.8 (0.38) | 0.4 (0.03) |
| Tier Op | 24.8 (0.51) | 20.9 (0.4) | 5.3 (0.19) | 0.4 (0.04) | 0.1 (0.01) | 73.4 (0.44) | 26 (0.51) |
| | | | | GOLD 1 & 2 | | | |
| Tier 0 | 41.8 (0.74) | 58.8 (0.85) | 23 (0.59) | 5.4 (0.41) | 0.5 (0.06) | 12.3 (0.49) | 34.8 (0.74) |
| Tier 1 | 25.7 (0.44) | 17.6 (0.64) | 50.2 (0.59) | 19.1 (0.64) | 5.9 (0.33) | 7.2 (0.28) | 25.8 (0.39) |
| Tier 2 | 9 (0.33) | 4.6 (0.39) | 28 (0.77) | 55.2 (0.9) | 11 (0.54) | 1.2 (0.14) | 13.3 (0.47) |
| Tier 3 | 6 (0.35) | 1.5 (0.2) | 19.4 (0.81) | 18.1 (0.78) | 59.2 (1.13) | 1.8 (0.25) | 7.7 (0.41) |
| Tier Op | 17.5 (0.52) | 14.5 (0.44) | 12.5 (0.41) | 0.9 (0.1) | 0.5 (0.06) | 71.6 (0.6) | 18.4 (0.5) |
| | | | | GOLD 3 & 4 | | | |
| Tier 0 | 12.2 (0.69) | 38.7 (1.34) | 35.4 (1.09) | 7.8 (0.7) | 1.3 (0.24) | 16.7 (1) | 10.1 (0.68) |
| Tier 1 | 28.1 (0.6) | 7.9 (0.63) | 50.3 (0.85) | 23.1 (0.92) | 11.2 (0.65) | 7.5 (0.44) | 25 (0.56) |
| Tier 2 | 21.5 (0.74) | 1.7 (0.3) | 16.5 (0.85) | 58.5 (1.06) | 22.3 (0.99) | 1 (0.13) | 22.8 (0.77) |
| Tier 3 | 24.9 (1.14) | 0.6 (0.26) | 8 (0.7) | 13.4 (0.98) | 76.8 (1.32) | 1 (0.15) | 29.7 (1.22) |
| Tier Op | 13.3 (0.72) | 7.9 (0.53) | 20.1 (0.67) | 1.6 (0.17) | 2.3 (0.23) | 68 (0.92) | 12.5 (0.6) |

Notes: Percent volumes for each tier at year 0 (left column) and 5 (right column), as well as the percent volume of tier reassignment over time interval (rows to columns). The Sankey plots in **Fig. 5** illustrate tier reassignment over 5 years. The flow lines are qualitative and not the actual quantitative values, which are provided here. All values are presented as means (standard error of the mean).

#### Supplemental Methods

**Clinical Metrics**: The collected data encompassed the age of subjects, their sex, body mass index (BMI), 6-minute walk distance, St. George's Respiratory Questionnaire (SGRQ), pack-years, smoking status (former/current), CT acquired lung volume at total lung capacity (TLC) and functional residual capacity (FRC), and pulmonary function test (PFT) results, which include forced vital capacity (FVC), forced expiratory volume in one second ($FEV_1$), and the ratio of $FEV_1$ to FVC (both as a percentage of the predicted value and the absolute value).

**High Resolution Computed Tomography**: For each participant at each phase, an inspiratory CT scan was performed during a breath hold at full inflation. Across scanners, the study maintains the same CT volumetric dose index that, for the Siemens Flash, translates to a mAs of 110, kV of 120, 0.5 s rotation speed, 0.625 mm slice thickness and 0.5 mm slice spacing. A full expiratory CT scan at relaxed deflation followed the same protocol at half the dose. There were dose adjustments for 3 levels of body mass indices. All CT data have been reconstructed using smooth kernels and have undergone extensive quality control (QC) to confirm fidelity. Scanner make, i.e., GE/Siemens/Philips, was also reported in this study.

**Spatial Alignment of Temporally Resolved Paired CT scans:** To evaluate changes in sub-volume tier assignment and location, we spatially aligned the Phase 2 PRM to the baseline PRM geometric frame. This was performed by 4D Medical. First, lungs were segmented from the thoracic cavity. Second, all inspiratory CT scans were spatially aligned to their paired expiratory CT scans within a single examination (i.e., Phase). Third, the expiratory CT scan at Phase 2 was spatially aligned to the Phase 1 expiratory CT scan. Finally, the transformation matrices for steps 2 and 3 were applied to the Phase 2 inspiratory CT scan. Once completed, all four CT scans shared the same geometric space. Corrections for differences in inflation levels were not performed.

**Parametric Response Map:** 3D PRM data were generated from all paired clinical CT data. In brief, lungs from paired CT scans (inspiration CT = Ins; expiration CT = Exp) were segmented from the thoracic cavity and spatially aligned to the geometric frame of the lungs captured on CT at full deflation as described above. Using in-house software, CT voxels, the smallest unit of volume on 3D imaging data, were classified based on paired Hounsfield Unit values (HU) as normal parenchyma ($PRM^{Norm}$; green; -950 ≤ Ins < -810 HU & -856 ≤ Exp < -250 HU), functional small airways disease ($PRM^{fSAD}$; yellow; -950 ≤ Ins < -810 HU & -1000 ≤ Exp < -856 HU), parenchymal disease ($PRM^{PD}$; purple; -810 ≤ Ins < -250 HU & -1000 ≤ Exp < -250 HU), and emphysema ($PRM^{Emph}$; red, -1000 ≤ Ins < -950 HU & -1000 ≤ & < -856 HU) [1-3]. The percent lung volumes of each PRM classification were calculated by normalizing the sum of all like-classed voxels by the total lung volume at deflation then multiplying by 100.

#### Supplemental Figure 2

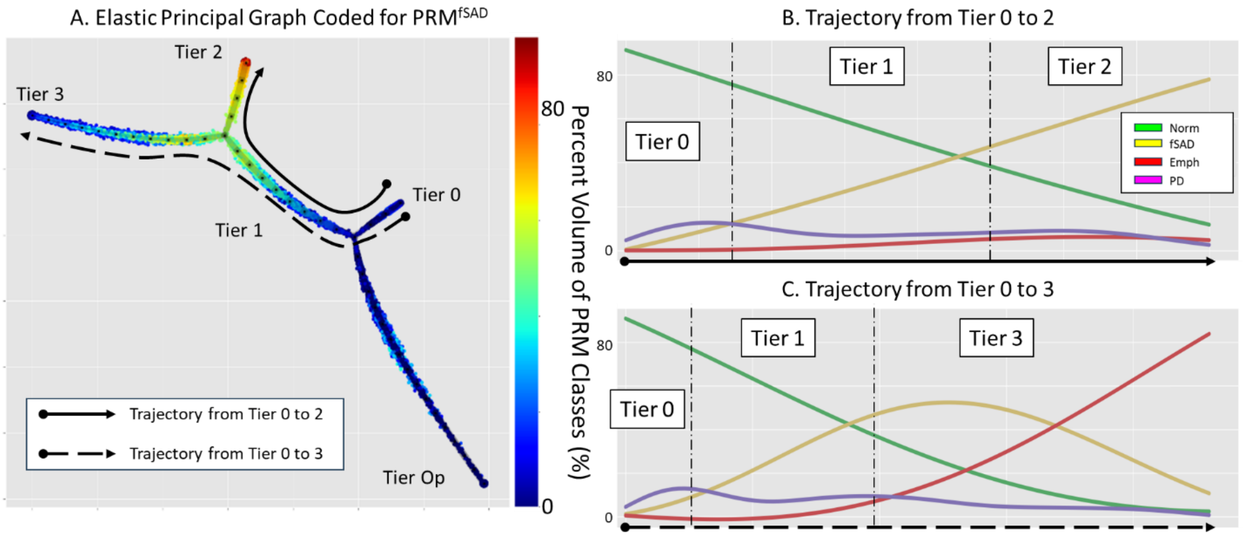


Profiles of PRM classifications infer progression along the elastic principal graph. (a) Markers on the elastic principal graph, representing sub-volumes from the 300 cases used to build the model, are color-coded by the percent volume of $PRM^{fSAD}$. Spline fits of the PRM values are provided for all four PRM classifications, Norm (green), fSAD (yellow), Emph (red) and PD (magenta), along the trajectories (b) Tier 0 to Tier 2 and (c) Tier 0 to Tier 3. Trajectories on the elastic principal graph are indicated as lines, solid for (b) and dashed for (c), with filled circle and arrowhead indicating beginning and end, respectively. Tiers along profiles in (b) and (c) are separated by dash-point lines.

In brief, the elastic principal graph constructs a low-dimensional approximation of data by embedding a graph whose nodes are positioned at the middle of the multidimensional data distribution, close to the data points. Simultaneously, it minimizes deviations from harmonic graph embedding, ensuring that the graph branches represent segments of smooth, nonlinear trajectories. Accordingly, data points can be partitioned accordingly to the proximity to the graph segments, and along each segment each data point can be characterized by a measure of pseudotime, which characterizes its relative position on the segment. In the simplest case, we construct a loopless principal graph, known as a principal tree [4].

#### References


1. Galbán, C.J., et al., *Parametric response mapping as an indicator of bronchiolitis obliterans syndrome after hematopoietic stem cell transplantation.* Biol Blood Marrow Transplant, 2014. **20**(10): p. 1592-8.
2. Belloli, E.A., et al., *Parametric Response Mapping as an Imaging Biomarker in Lung Transplant Recipients.* Am J Respir Crit Care Med, 2017. **195**(7): p. 942-952.
3. Galbán, C.J., et al., *Computed tomography-based biomarker provides unique signature for diagnosis of COPD phenotypes and disease progression.* Nat Med, 2012. **18**(11): p. 1711-5.
4. Golovenkin, S.E., et al., *Trajectories, bifurcations, and pseudo-time in large clinical datasets: applications to myocardial infarction and diabetes data.* Gigascience, 2020. **9**(11).
